# A community based cross-sectional study on impact assessment of COVID-19 on mental health in Central India

**DOI:** 10.1101/2023.08.08.23293808

**Authors:** Dharmendra Gahwai, Sonal Dayama, Ravindra Kumar, Sandip Kumar Chandraker, Akanksha Mishra, Babita Sahu, Mini Sharma

## Abstract

The current study is one of few community based surveys assessing the effect of COVID -19 in rural India. It is a cross sectional study to find the prevalence of depression, anxiety and PTSD among those affected with COVID-19 disease. Generalized Anxiety Disorder Assessment (GAD-7), PHQ-9 and IES-R were used to assess prevalence levels of depression, anxiety, and PTSD among the community of Durg district of Chhattisgarh state of India. Out of total 431 participants, 44 % were male, mean age of participants was 41±14, 87% had health insurance, 40% had co-morbidities like hypertension or diabetes, half of them experienced food shortage and change in income during the pandemic. One third participants experienced death among one or more family members due to the pandemic. The mean scores of IES-R, GAD-7 and PHQ-9 were 23.59-24.91, 1.50,2.07 and 1.06-1.58 respectively. Thirty percent observed some distress, 15% reported depression and 12 % reported anxiety. The adjusted effect of death in family due to COVID-19 was found to be significantly associated with higher risk of mental distress, whereas education was associated with lower risk of distress. Depression and anxiety was more among the elderly and less among individuals living in nuclear families. Scarcity of food and change in income were significantly associated with anxiety.

## Introduction

The current literature suggests that people affected by COVID-19 may have a high burden of mental health problems, including depression, anxiety disorders, stress, panic attack, irrational anger, impulsivity, somatization disorder, sleep disorders, emotional disturbance, posttraumatic stress symptoms, and suicidal behaviors.(1)(2) (3) (4) A WHO press release in March 2022, said that global prevalence of anxiety and depression increased by a massive 25% in the first year of the COVID-19 pandemic.(5)

A systematic review conducted during initial phase of the pandemic revealed relatively high rates of symptoms of anxiety (6.33% to 50.9%), depression (14.6% to 48.3%), post-traumatic stress disorder (7% to 53.8%), psychological distress (34.43% to 38%), and stress (8.1% to 81.9%) are reported in the general population during the COVID-19 pandemic in China, Spain, Italy, Iran, the US, Turkey, Nepal, and Denmark. (6) In a recent published online survey of participants of 40 countries, probable depression was detected in 17.80% and distress in 16.71%.(7)

A recently published systematic review and meta-analysis indicated the prevalence of delirium as a common occurrence among patients hospitalized due to severe coronavirus infections (severe acute respiratory syndrome (SARS-CoV) and Middle East respiratory syndrome (MERS)), whereas post-traumatic stress disorder (PTSD), anxiety, depression, and fatigue were observed in the subsequent months. (8)

Many studies were conducted during the onset of the pandemic to identify stress, anxiety and depression and PTSD among specific populations like Health Care workers, students, adolescents, patients of mental diseases etc. There has been extensive research on the COVID-19 disease and more are still being conducted. Researchers are conducting live systematic reviews on mental health related to COVID-19. (9) Chhattisgarh, a state in India, has one of the highest suicide rates (26.4%) (The national average is 11.3%).(10) It was the twelfth highest affected state of the COVID-19 pandemic in India.(11) Very few were conducted among general population in India. The current study is one of few community based surveys conducted in rural India. It is a cross sectional study to find the prevalence of depression, anxiety and PTSD among those affected with COVID-19 disease in past three years.

## Methodology

A community based survey was conducted among rural and urban blocks of district Durg of the state of Chhattisgarh. The district is divided into four rural and one urban block. All five blocks were visited. Four villages in the practice areas of two PHCs in rural and Urban PHC and CHC in urban area of each block were conveniently selected. Out of these 18 villages/urban ward, house visits facilitated by the health care workers were undertaken. Those houses were any one member of the family (adult or child) who was either dead or diagnosed with COVID-19 any time, were selected and a senior family member (adult) who was living with the person affected was interviewed.

Sample size Estimation: With confidence level of 95%, assuming 50% prevalence from studies cited above, sample size estimated was 384. Considering non response rate of 10%, sample size estimated was 422. During the course of the study, we interviewed for 435 participants and included data of 431 for analysis.

Survey instrument consisted of two sections. One section was on sociodemographic data (age, gender, marital status, educational level, number of siblings, occupation, employment, monthly family income) of the participant and his/ her family and medical history of the one who had been affected with COVID-19 included date of diagnosis, relevant personal history, hospital admissions or home isolation, presence of complications and health problems related to the disease. Those with preexisting mental illness (diagnosed before March 2020) were excluded from the study.

For study purposes affected by COVID-19 meant, getting diagnosed with COVID-19 and getting treated at home or hospital.

The second section consisted of psychometric assessment scales namely the PHQ 9, GAD 7 & IES-R scale.

1. The Generalized Anxiety Disorder Assessment (GAD-7) is a seven-item instrument that is used to measure or assess the severity of generalized anxiety disorder (GAD). Each item asks the individual to rate the severity of his or her symptoms over the past two weeks. Response options include “not at all”, “several days”, “more than half the days” and “nearly every day”. It has been has been validated for primary care patients, general population, and adolescents with GAD. (12)
2. The PHQ-9 is the nine item depression scale of the Patient Health Questionnaire. The PHQ-9 is a useful tool in screening for depression in primary care. (13) Certified Hindi translation of the tools PHQ-9 and GAD-7 are available on the official website of the authors. (14)
3. Impact of Event Scale - Revised (IES-R) is a 22-item self-report measure that assesses subjective distress caused by traumatic events. Respondents are asked to identify a specific stressful life event and then indicate how much they were distressed or bothered during the past seven days by each "difficulty" listed (15).

Ethical aspects: The study was approved by Institutional Ethics Committee of National Institute of Research in Tribal Health (NIRTH), Indian Council of Medical Research, Jabalpur, Madhya Pradesh & IEC of Pt. JNM Medical College, Raipur.

### Statistical Analysis

A cutoff score of L4, L4, L15 was used to determine the presence of depression, anxiety and stress respectively. Tables were used to present the findings. Frequency (percentage) and mean ± sd were used to describe the data.

First, we applied chi-square test to see univariate association of different covariates with IES-R, PHQ-9 and GAD-7 scores. Then, we estimated logistic regression model to assess adjusted association. The covariates for which the univariate association were significant were considered in logistic regression model. As, only single variable for PHQ-9 and two variables for GAD-7 were significant in Chi-square test, so we did not run logistic regression for these mental health indicators. A p-value of less than 0.05 was considered statistically significant. All the analyses was done using IBM SPSS version 26.

## Results

During the survey, a total of 435 patients were recruited and interviewed. Out of which some information was missing for four individuals hence was excluded from the analysis. Therefore, analysis of the results was performed 431 responses. The mean age of participants was 41±14 years. There were 190 (44.1%) male and 241 (55.9%) female. Out of these 431 participants, only 117 (27.15%) were unemployed and rest were engaged in active work. Other demographic details are mentioned in table 1. Co-morbidity in terms of hypertension (20.9%), diabetes (11.1%), Thyroid disorders (3%), CAD (2.1%), Others (7.2%) were observed. As expected, comorbidity was significantly associated with the older age (p<0.001). Most of the participants (69.1%) had less than 6 members in their family. Median monthly family income was Rs.15000 (min=1000, max=180,000), 63.3% live in rural areas, 19.7% were graduate and 26.9% had attained education only up to primary level.

1. **IES-R scale-**A total of 113 (26.2%, 43 males (22.6%) and 70 (29.0%) females) participants had symptoms concerned with PTSD. Forty-five (10.4%) had scores 33 and above. Married person had approximately 40 percent lower risk of distress (OR=0.590, CI: 0.368-0.948, p-value=0.032). Elderly person was more prone to distress (p-value =0.014). Alcohol consumption and habit of smoking had lowered the risk of distress by 44 percent (OR=0.559, CI:(0.331,0.943) and 54 percent (OR=0.460, CI:(0.256,0.824), P-value=0.028) respectively. Presence of comorbidity and death in family due to Covid-19 exacerbated the traumatic distress (Table 2). Higher education level was associated with increased risk of facing stress (p-value=0.018) while unemployed individual was less prone to distress (p-value=0.029). Out of pocket expenditure for medical expenses had significant toll on mental distress (Table 2). Multinomial logistic regression analysis was performed with the variables found to be statistically significant in univariate analysis (Chi-square test of association) to see the significance of adjusted effect while controlling for remaining factors. The fitted multinomial logistic model was significant (Table 3). The goodness of fit test failed to reject the null hypothesis that the model fits the data well (p = 0.744). The results had shown that lower level of education (up to primary level) was associated with lower risk of distress. The adjusted effect of death in family due to COVID-19 was also found to be significantly associated with higher risk of mental distress (Table 3).
2. **PHQ-9 scale**- As per PHQ-9 score, total 51(11.8%) participants were facing problem of depression out of which 20 (10.5%) were males and 31 (12.9%) were females. Out of these, 46 had mild depression and 4 had moderate and 1 had severe depression. Chi-square test for association had shown that age-group, scarcity of food and change in income were significantly associated with depression (Table 2). Older individuals were more likely of facing depression. Individuals living in nuclear families reported less depression.
3. **GAD-7 scale**-On administering this instrument, a total 64 (14.8%) participants were found abnormal in which 25 (13.2%) were male and 39 (16.2%) were female. Fifty three had mild anxiety, seven had moderate and four had severe anxiety. Chi-square test for association revealed that age group, education level, scarcity of food and change in income were significantly associated with anxiety (Table 2). Participants with higher level of education and older age were more prone of being anxious.

**Table 1:** Sociodemographic profile of subjects interviewed.

|  | N=431 | Frequency (%) |
| --- | --- | --- |
| Age ( years) | 18-30 | 124(28.8) |
|  | 31-60 | 262(60.8) |
|  | >60 | 45(10.4) |
| Sex | Male | 190(44.1) |
|  | Female | 241(55.9) |
| Marital Status | Married | 323(74.9) |
|  | Unmarried | 108(25.1) |
| Occupation | Skilled/Semi-professional/Professional | 138(32.0) |
|  | Semi-skilled/shop-owner/farmer/student | 176(40.8) |
|  | Unemployed | 117(27.1) |
| Education level | Up to primary level | 85(19.7) |
|  | Middle/high/higher | 230(53.4) |
|  | Graduate/Professional | 116(26.9) |
| Change in Income due to Covid-19 | No change | 210(48.7) |
|  | Decreased | 221(51.3) |
| Food Shortage during Covid-19 | Never | 66(15.3) |
|  | Sometime | 94(21.8) |
|  | Often | 271(62.9) |
| Comorbidity | Absent | 259(60.1) |
|  | Present | 172(39.9) |
| Type of Family | Nuclear | 262(60.8) |
|  | Joint | 189(39.2) |
| Health Insurance | Yes | 375(87.0) |
|  | No | 56(13.0) |
| Substance Abuse | Yes | 172(39.9) |
|  | No | 259(60.1) |
| Type of Substance Abuse | Alcohol | 118(27.4) |
|  | Smoking | 100(23.2) |
|  | Tobacco | 86(20.0) |
|  | Others | 2(0.5) |
| Physical Activity | Yes | 115(26.7) |
|  | No | 316(73.3) |
| Duration of exercise | 30 to 45 min | 83(19.3) |
|  | more than 45 | 31(7.2) |
|  | < 30 or none | 317(73.5) |
| Food preference | Veg | 97(22.5) |
|  | Non-veg | 334(77.5) |
| Year in which affected | 2020 | 54(12.5) |
|  | 2021 | 370(85.8) |
|  | 2022 | 7(1.6) |
| Family member died due to Covid-19 | Yes | 131(30.4) |
|  | No | 300(69.6) |
| Relationship with | Parents/Spouse/Siblings/Child | 106(80.9) |
| deceased |  |  |
|  | Others | 25(19.1) |
| Time of death | Less than one year | 6(4.6) |
|  | More than one year | 125(95.4) |
| Mental stress presence | Combination of result of all scales (at least one of the three scales have shown presence of mental health issue) | 157(36.4) |
|  | IES-R | 113(26.2) |
|  | PHQ-9 | 51(11.8) |
|  | GAD-7 | 64(14.8) |
| IES-R | None | 318(73.8) |
|  | Mild | 69(16.0) |
|  | Moderate | 13(3.0) |
|  | Severe | 31(7.2) |
| GAD-7 | None | 367(85.2) |
|  | Mild | 53(12.3) |
|  | Moderate | 7(1.6) |
|  | Severe | 4(0.9) |
| PHQ-9 | None | 380(88.2) |
|  | Mild | 45(10.4) |
|  | Moderate | 5(1.2) |
|  | Severe | 1(0.2) |
| Average score (95% CI) | IES-R | 24.25(23.59-24.91) |
|  | GAD=7 | 1.78(1.50,2.07) |
|  | PHQ-9 | 1.32(1.06-1.58) |

**Table 2:** Association of Mental Health with socio-demographic factors.

| **Factors** |  | **IES-R (% or 95% C.I)** | **PHQ-9 (% or 95% C.I)** | **GAD-7 (% or 95% C.I)** |
| --- | --- | --- | --- | --- |
| Age group | 18-30 | 21(16.9) | 5(9.8) | 9(7.3) |
|  | 31-60 | 76(29.0) | 36(13.7) | 46(17.6) |
|  | > 60 | 16(35.6) | 10(22.2) | 9(20.0) |
|  | P-value | **0.014** | **0.001** | **0.011** |
| Gender | Male | 43 (22.6) | 20 (10.5) | 25 (13.2) |
|  | Female | 70 (29.0) | 31 (12.9) | 39 (16.2) |
|  | Odds Ratio | 1.399 (0.902-2.171) | 1.255 (0.690-2.280) | 1.274 (0.741-2.193) |
|  | P-value | 0.152 | 0.548 | 0.415 |
| Marital Status | Married | 76 (23.5) | 40 (12.4) | 46 (14.2) |
|  | Single | 37 (34.3) | 11 (10.2) | 18 (16.7) |
|  | Odds ratio | **0.590 (0.368-0.948)** | 1.246 (0.615-2.525) | 0.831 (0.458-1.504) |
|  | P-value | 0.032 | 0.609 | 0.535 |
| Education | Up to primary | 23 (19.8) | 9 (7.8) | 11 (9.5) |
|  | Middle to higher | 58 (25.2) | 29 (12.6) | 34 (14.8) |
|  | Graduate | 32 (37.6) | 13 (15.3) | 19 (22.4) |
|  | P-value | **0.018** | 0.212 | **0.042** |
| Employment/<br>Occupation | Skilled | 34 (24.5) | 20 (14.5) | 19 (13.8) |
|  | Semi-skilled | 57 (32.4) | 22 (12.5) | 29 (16.5) |
|  | Unemployed | 22 (18.8) | 9 (7.7) | 16 (13.7) |
|  | P-value | **0.029** | 0.211 | 0.734 |
| Type of family | Nuclear | 62 (23.7) | 24 (9.2) | 37 (14.1) |
|  | Joint | 51 (32.2) | 27 (16.0) | 27 (16.0) |
|  | Odds ratio | 0.717 (0.464-1.108) | **0.530 (0.295-0.955)** | 0.865 (0.505-1.481) |
|  | P-value | 0.145 | 0.046 | 0.677 |
| Type of house | Own | 110 (26.1) | 50 (11.9) | 63 (15.0) |
|  | Rental | 3 (30.0) | 1 (10.0) | 1 (10.0) |
|  | Odds ratio | 0.825 (0.210-3.247) | 1.213 (0.150-9.777) | 1.585 (0.197-12.658) |
|  | P-value | 0.726 | 1.000 | 1.000 |
| Family income | Up to 10000 | 47 (32.2) | 20 (13.7) | 26 (17.8) |
|  | 10001-20000 | 34 (22.2) | 19 (12.4) | 20 (13.9) |
|  | Above 20000 | 32 (24.2) | 12 (9.1) | 18 (13.6) |
|  | P-value | 0.125 | 0.463 | 0.469 |
| BPL card holder | Yes | 82(25.2) | 36(11.1) | 52(16.0) |
|  | No | 31(29.2) | 15(14.2) | 12(11.3) |
|  | Odds ratio | 0.816(0.501-1.330) | 0.756(0.396-1.443) | 1.492(0.763-2.916) |
|  | P-value | 0.446 | 0.390 | 0.273 |
| Public Health Insurance | Yes | 98(26.1) | 40 (10.7) | 54 (14.4) |
|  | No | 15(26.8) | 11 (19.6) | 10 (17.9) |
|  | Odds ratio | 0.967(0.513-1.824) | 0.488(0.234-1.020) | 0.774(0.368-1.626) |
|  | P-value | 1.000 | 0.073 | 0.545 |
| Alcohol | Yes | 22(18.6) | 11(9.3) | 19(16.1) |
|  | No | 91(29.1) | 40(12.8) | 45(14.3) |
|  | Odds ratio | **0.559 (0.331-0.943)** | 0.702(0.347-1.418) | 1.143(0.638-2.049) |
|  | P-value | 0.028 | 0.404 | 0.651 |
| Smoking | Yes | 16 (16.0) | 7 (7.0) | 15 (15.0) |
|  | No | 97 (29.3) | 44 (13.3) | 49 (14.8) |
|  | Odds ratio | 0.460 (0.256-0.824) | 0.491(0.214-1.127) | 1.016 (0.542-1.901) |
|  | P-value | 0.009 | 0.111 | 1.000 |
| Tobacco | Yes | 23(26.7) | 12(14.0) | 16(18.6) |
|  | No | 90(26.1) | 39(11.3) | 48(13.9) |
|  | Odds ratio | 1.034(0.606-1.767) | 1.272(0.635-2.551) | 1.414(0.759-2.637) |
|  | P-value | 0.892 | 0.462 | 0.309 |
| Comorbidity | Yes | 54(47.8) | 17(9.9) | 31(18.0) |
|  | No | 59(22.8) | 34(13.1) | 33(12.7) |
|  | Odds ratio | **1.551(1.005-2.393)** | 0.726(0.392-1.345) | 1.506 (0.883-2.564) |
|  | P-value | 0.048 | 0.362 | 0.166 |
| Casualty/Death in Family due to Covid-19 | Yes | 64(49.2) | 17(13.1) | 25(19.2) |
|  | No | 49(16.3) | 34(11.3) | 39(13.0) |
|  | Odds ratio | **4.975 (3.145-7.874)** | 1.182 (0.634-2.203) | 1.600 (0.922-2.775) |
|  | P-value | <0.001 | 0.627 | 0.105 |
| Out of pocket expenditure (Rs.) | Nil | 14(20.6) | 8(11.8) | 6(8.8) |
|  | Up to 5000 | 24(14.5) | 13(7.9) | 23(13.9) |
|  | 5000-10000 | 10(25.6) | 4(10.3) | 4(10.3) |
|  | Above 10000 | 64(40.8) | 26(16.6) | 31(19.7) |
|  | P-value | **<0.001** | 0.117 | 0.122 |
| Rural/Urban | Rural | 69(25.3) | 32(11.7) | 41(15.0) |
|  | Urban | 44(27.8) | 19(12.0) | 23(14.6) |
|  | Odds ratio | 0.876(0.563-1.364) | 0.971(0.531-1.779) | 1.037(0.597-1.802) |
|  | P-value | 0.571 | 1.000 | 1.000 |
| Caste | General | **14(34.1)** | 7(17.1) | 9(22.0) |
|  | OBC | **70(25.3)** | 29(10.5) | 35(12.6) |
|  | SC | **20(25.6)** | 9(11.5) | 13(16.7) |
|  | ST | **8(23.5)** | 6(17.6) | 7(20.6) |
|  | P-value | **0.659** | 0.444 | 0.285 |
| Food scarcity | Often | 24(39.3) | 9(14.8) | 14(23.0) |
|  | Sometime | 28(26.7) | 20(19.0) | 19(18.1) |
|  | Never | 61(23.0) | 22(8.3) | 31(11.7) |
|  | P-value | **0.033** | 0.012 | 0.047 |
| Change in income | Decreased | 65(29.4) | 34(15.4) | 43(19.5) |
|  | No change | 48(22.9) | 17(8.1) | 21(10.0) |
|  | Odds ratio | 1.406(0.912,2.169) | 2.066(1.115,3.817) | 2.174(1.241-3.808) |
|  | P-value | 0.127 | 0.025 | 0.007 |
| Health insurance | Yes | 98(26.1) | 40(10.7) | 54(14.4) |
|  | No | 15(26.8) | 11(19.6) | 10(17.9) |
|  | Odds ratio | 0.967(0.513-1.824) | 0.488(0.234-1.020) | 0.774(0.368-1.626) |
|  | P-value | 1.000 | 0.073 | 0.545 |

**Table 3:** Result of multinomial logistic regression.

| IES-R Status<sup>a</sup> | P value | OR (95%CI) |
| --- | --- | --- |
| Age > 60 | 0.322 | 1.636 (0.618-4.331) |
| Age 31-60 | 0.184 | 1.544(0.813-2.933) |
| Age 18-30 | . | . |
| OOPE (>10000) | 0.113 | 1.827(0.867-3.849) |
| OOPE (5001-10000) | 0.927 | 1.048(0.379-2.900) |
| OOPE (Up to 5000) | 0.156 | 0.563(0.254-1.246) |
| OOPE (Nil) | . | . |
| Skilled | 0.091 | 1.863(0.905-3.835) |
| Semi-skilled | 0.181 | 1.591(0.805-3.144) |
| Unemployed | . | . |
| Education (middle to higher) | 0.256 | 0.669(0.334-1.338) |
| Education (up to primary) | 0.053 | 0.430(0.182-1.011) |
| Education (Graduate) | . | . |
| Smoking (No) | 0.356 | 1.635(0.575-4.646) |
| Smoking (Yes) | . | . |
| Married | 0.645 | 0.871(0.483-1.570) |
| Single | . | . |
| Alcohol (No) | 0.481 | 1.409(0.543-3.655) |
| Alcohol (Yes) | . | . |
| Comorbidity (Present) | 0.449 | 1.228(0.722-2.089) |
| Comorbidity (Absent) | . | . |
| Casualty in family due to Covid-19 disease (No) | 0.000 | **0.262(0.157-0.437)** |
| Casualty in family due to Covid-19 disease (Yes) | . | . |
| Shortage of food during pandemic (never) | 0.039 | 0.463(0.223-0.964) |
| Shortage of food during pandemic (sometimes) | 0.131 | 0.549(0.252-1.195) |
| Shortage of food during pandemic (often) | . | . |
| a. The reference category is: Normal. |  |  |
| Pseudo R-Square = 0.275, p-value (Goodness-of-Fit) = 0.612 |  |  |
| p-value (final model vs. Intercept only) =0.000 |  |  |

## Discussion

Proportion of participants in the current study having symptoms of PTSD, anxiety and depression was 10.4%, 11.8% and 14% respectively. Similar rates of depression and post-traumatic-stress disorders were found in 6% and 10% respectively one year after infection in one year cohort study in Austria. (16)

Relatively high rates of symptoms of anxiety (6.33% to 50.9%), depression (14.6% to 48.3%), post-traumatic stress disorder (7% to 53.8%), had been reported in the general population during the COVID-19 pandemic in China, Spain, Italy, Iran, the US, Turkey, Nepal, and Denmark. (6)

A systematic review and meta-analysis conducted in the early phase (2020) of the pandemic, of 66 studies with 221,970 participants, reported the overall pooled prevalence of depression, anxiety, distress, and insomnia was 31.4%, 31.9%, 41.1% and 37.9%, respectively. Noninfectious chronic disease patients, quarantined persons, and COVID-19 patients had a higher risk of depression (Q=26.73, p<0.01) and anxiety (Q=21.86, p<0.01) than other populations.(17)

In 2021, another systematic review conducted among 40 out of 167 developing countries in Africa, Asia (East, Southeast, South, and West), Europe, and Latin America, summarized the prevalence rates of mental health symptoms based on 341 empirical studies with a total of 1,704,072 participants. Distress (29%) and depression (27%) were the most prevalent.(18)

While high rates of Anxiety and Depression (73% & 70%) among Bosnia and Herzegovina Citizens were reported during the third Wave of COVID-19, our study reported quite the opposite. (19)

In the present study, old age group were more likely facing depression. Nuclear families reported less depression. Participants with higher level of education and older age were more prone of anxiety in the present study. While in some other studies, factors were associated with depression and/or anxiety were living alone (20) lower educational level (21) (22) higher education level (23) living in urban areas (21), (24), or in rural areas (25),(20), female gender (6),(26), (27), (22), (28), while reports on age as a risk factor were varying in some other studies (6)(21,22) (29). A study conducted among Malaysia population during the third wave of the pandemic reported age, gender, and friends infected with virus were the three important predictors of depression and anxiety. The odds of having depression (OR = 1.44; C·I. = 1.32- 1.62) and anxiety (OR = 1.36; C·I. = 1.27-1.47) were significantly higher among females than in males. (30) In our study, gender was not found to be a significant risk factor, through older age group was significantly associated with depression and anxiety.

In the present study, we found that marriage, young age group, alcohol and smoking consumption, unemployment had lower risk of PTSD in univariate analysis and co-morbidity and death in the family due to COVID-19 increased the risk of PTSD. In multivariate analysis lower level of education (up to primary level) was associated with lower risk of distress while, death in family due to COVID-19 was also found to be significantly associated with higher risk of PTSD. A review of 33 studies of 6743 participants concluded that the long-term effect from direct COVID-19 infection has been associated with no or mild symptoms. Studies exhibited the long-term prevalence of anxiety, depression, PTSD, and sleep disturbances to be comparable to general population levels suggesting deterioration could be attributed to indirect effects of COVID-19 psychosocial factors. (31) Presence of chronic/psychiatric illnesses, unemployment were found significantly associated with anxiety, depression many studies. (6)

The loss of a family member or friend due to direct effect of a pandemic like SARS-COV2 infection amplifies psychological distress (32)(33,34). Many studies report increased incidence major depressive episodes, panic disorder, and post-traumatic stress disorder after unexpected death.

Our study presents some limitations. The information about co-morbidities was self-reported. The association of intensity or severity of a mental health issue with closeness / friendliness with the type (like sibling, spouse, etc) of relative who succumbed to the pandemic has not been analyzed. Severity of disease like getting hospitalized, being on ventilator and near death experiences may also impact the mental status. This has not been elicited in our study.

Social factors which may have influence on mental health status like income, financial/ Economic burden, food scarcity and stigma has not been captured in detail through any standard tool.

As highlighted there were many studies on the impact of Covid-19 on mental health during the first wave of the pandemic. With the progress of pandemic, as scientific studies found more about the disease, the global perception and attitude about the virus changed. We assume that this might have changed the mental health impact as well. This is evident in our results. The proportion of depression, anxiety and PTSD is lowest or similar to the studies discussed above. One of the strengths of the present study is that this is that is it one of very few in- person community based studies among general population, who had at least one close family member affected (infected or died) with the disease. Most of the studies discussed are either conducted among special groups like mothers, health care providers, students, professionals or conducted through online surveys.

Conclusion: Scarcity of food and decrease in income and increasing age were found to be significantly associated with anxiety and depression among the participants. In future, governments should remember to include social security measures as an integral part of pandemic readiness.

Death in family due to COVID-19 was found to be significantly associated with higher risk of mental distress. We conclude that high proportion of PTSD found in our study population beckons, mental health physician and policy makers for mental health preparedness in future pandemics.

### Ethical considerations

The study was approved by Institute Ethics Committee of Pt. JNM Medical College, Raipur, Chhattisgarh.

## Highlights of the study

- Community based study in central India
- Anxiety and depression associated with scarcity in food and change in income
- Post -traumatic Stress Disorder post Covid-19 pandemic associated with death in family

### Funding Source

The study was financially supported under the intramural research scheme of Model Rural Health Research Unit of Department of Health Research, New Delhi, Government of India.

### Contributors

DG, RK & BS conceptualized the study. SD, BS and MS did the literature search and design the study. SD and AM were instrumental in acquisition of data. RK, SD and SC conducted analysis and interpretation of data. SD and RK drafted the article. MS, SD and RK revised the manuscript for important intellectual content. All authors approved the final approval of the version of the manuscript.

## Data Availability

Data can be made available on reasonable request

## Acknowledgement

The authors acknowledge Secretary, Department of Health Research and Director General, ICMR, New Delhi for providing facilities. Authors are thankful to Director, ICMR-NIRTH, Jabalpur for guidance and kind support in fructification of this project. We also acknowledge the cooperation of the participants and the support of the field functionaries of the Health and the Women & Child Welfare departments in facilitating the interviews. We thank the support staff of MRHRU, Jheet and Statistician (Mr Shubham Mishra) for support in data cleaning and analysis.

